# Phylogeogrphy reveals expansion of yellow fever virus genotypes in West and Central Africa

**DOI:** 10.1101/2023.06.06.23291042

**Authors:** Babatunde Olanrewaju Motayo, Adewale Opayele, Paul Akiniyi Akinduti, Adedayo Omotayo Faneye, Isibor Patrick Omoregie, Solomon Uche Oranusi

## Abstract

Yellow Fever virus remains the most medically significant Arbovirus in Africa, with the occurrence of large outbreaks with human fatalities in recent years in Africa. Molecular epidemiology has shown the presence of 4 genotypes circulating across Africa; however, paucity of data still exists regarding directional spread and phylogeography of the African Yellow fever genotypes. The need to fill this gap with information from spatiotemporal data from continuous occurrence of YF outbreaks in Africa conceptualized this study; which aims to investigate the most recent transmission events and directional spread of YF virus using updated genomic sequence data. Archived Yellow Fever sequence data was utilized along with epidemiologic data from outbreaks in Africa, to analyze the case/fatality distribution. Phylogeograhic analysis was also utilized to demonstrate the ancestral introduction and geographic clustering of YF genotypes in Africa. Directional spread and geographic transmission of YF was also investigated. African YF genotypes were found to be geographical distinct, circulating within distinct geographical boundaries. Spatiotemporal spread however revealed an eastward spread of the West African genotypes over time, and recent northward movement of the East African genotype. We conclude by recommending expanded human/ vectoral surveillance of YF and other Arboviruses of public health importance, and upscaling sequencing capabilities of new and existing public health labs in Africa to help in the defense against public health threats.

## INTRODUCTION

Yellow fever virus is the causative agent for yellow fever, a viral illness that affects mainly the liver, and may cause serious complications including hemorrhage and death [1]. It is a member of the family Flaviviridea, genus flavavirus, along with Dengue virus, West Nile virus and Japanese encephalitis virus. It is an enveloped virus with a single stranded positive-sense RNA genome of 11kb size, that encodes a single polyprotein that is cleaved into 3 structural proteins, envelope (E), matrix (M), and capsid proteins (C) and 7 non-structural proteins, NS1, NS2A, NS2B, NS3, NS4A, NS4B, and NS5 [2]. Yellow fever is an arthropod borne infection, maintained by the mosquito vector *Aedes aegypti*, and *Aedes albopticus*, although several vectorial studies have identified YF virus in other mosquitoes’ species, such as Aedes *taylori* [3]. The main vector of the YF virus is also capable of transmitting other viral infections such as Chikungunya (CHIK), Dengue (DENV) and Zika (ZIKV) in tropical and sub-tropical areas [3].

There are two major vectorial cycles of YF, the sylvatic also known as Jungle YF maintained in non-human primates (NHP) by the mosquito *Heamgogus spp* [4], the second cycle is the urban cycle where humans are the primary host and are transmitted by the peri urban mosquito *Aedes aegypti* [3].

Yellow fever virus has one serotype, but phylogenetic analysis has identified up to 5 genotypes West Africa genotype I, East Africa genotype, East Africa/Angolan genotype, South American genotype I and South American genotype II [5]. Previous genetic analysis has shown that these genotypes are geographically defined [6], as have been reported for other viruses [7–9]. Previous reports have also shown the likelihood of YF virus originating from around East Africa before emerging in West Africa. Also, majority of the studies carried out in Africa are directed towards outbreak investigation and genotype identification [6], only very few studies have attempted to investigate spatio temporal dynamics of YF in Africa [6, 10]. In a previous study West African YF sequences were shown to be more divergent with evidence of adaptive evolution to the sylvatic environment [10]. In another study, the phylogeography of YF showed stable population demography, regional confinement of genotypes and zoonotic spillovers from the sylvatic outbreaks in Africa [11]. Yellow fever outbreaks continue to occur in Africa despite availability of vaccine and mass deployment to at risk regions in various African countries [12]. For instance, between 2017 to 2022 there have been outbreaks of YF in 4 different states in Nigeria, with a total of 287 confirmed cases [13,14]. Outbreaks have also been reported in Senegal in 2020 with 542 suspected cases and 15 laboratory confirmed cases [15]. There have also been silent outbreaks in Nigeria, with a report among patients presenting with pyrexia of unknown origin [16].

The continuous reports of outbreaks of YF virus in different parts of Africa, despite implementation of mass vaccination campaigns calls for attention and motivated the conceptualization of this study. The current study was designed to investigate the relationship between YF field surveillance data and available genomic data to the continuous occurrence of outbreaks in Africa; as well as most recent geographic transmission events of Yellow Fever virus genotypes using updated sequence data.

## METHODS

### Data curation

Data relating to all reported outbreaks of yellow fever virus infections were collected after database search. The databases that were searched include PubMed, Scopus and Google Scholar. Supplementary Table 1 shows the list of Outbreaks with the corresponding articles they were reported in. Figure 1 shows a world map and with an enlarged map of Africa, with the names of countries labelled within each country’s geographical locations.

**Figure 1.**
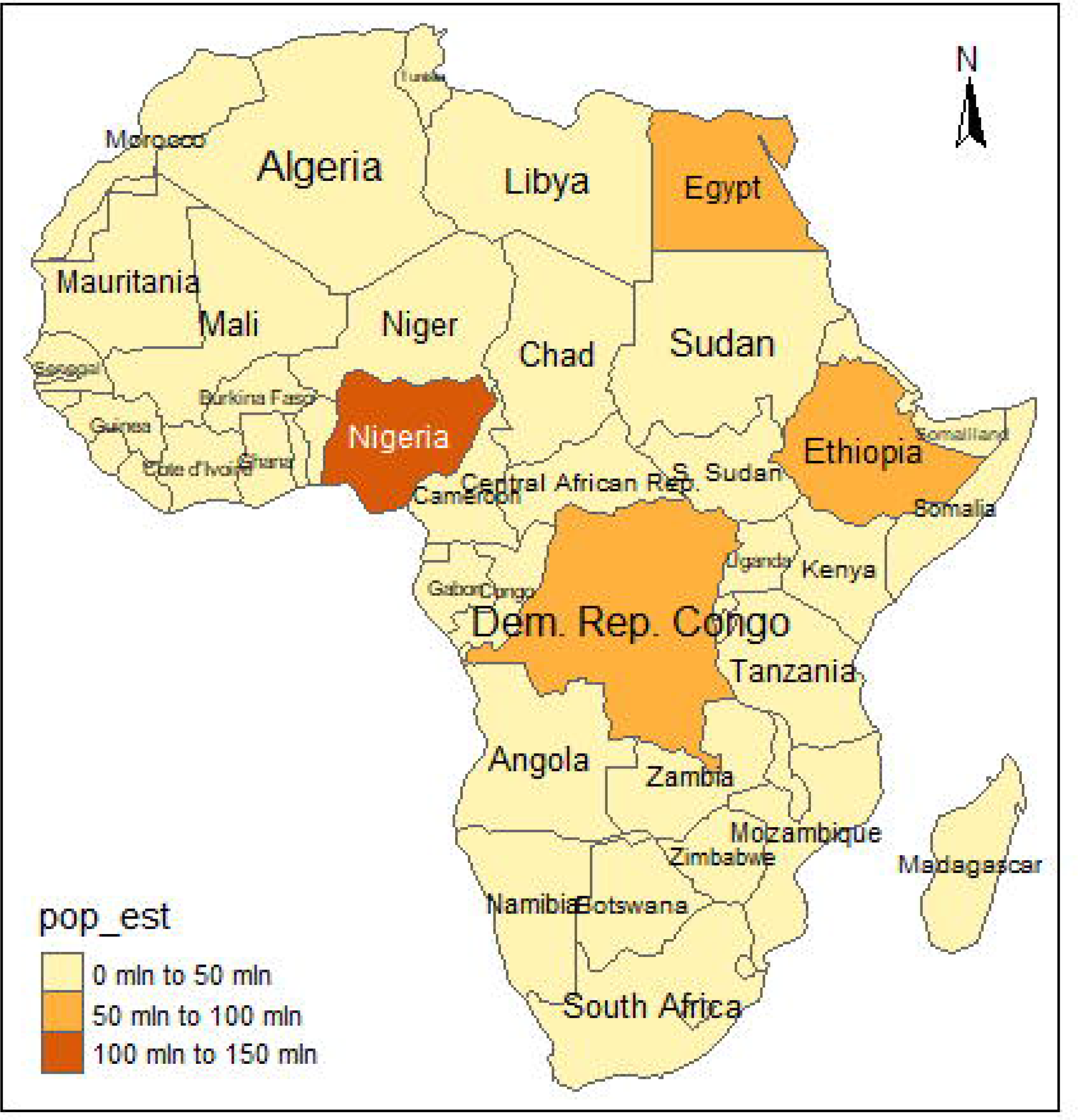
World map with various continents in color, and a map of Africa showing the different countries. The legend of the world map shows the color code for continents.

Partial YF, E gene sequences from Africa were downloaded from GenBank, along with sequences that were recovered from returnee traveler from Angola to China after the 2016 Angolan YF outbreak that spread to China via international travel [17]. This was done with the help of the NCBI Virus database. The dataset consisted of genome sequences of all archived E gene sequences from Africa, along with whole genome sequences deposited in GenBank, the E gene was selected because it represents the most frequently and consistently sequenced gene region for YF virus. In addition to the sequence data, metadata was also extracted such as sample source, year of isolation, and Country Location of isolation.

### Demographic analysis

The number of reported cases and subsequent deaths of infected individuals according to Country and year, were retrieved from the published reports of YF outbreaks in Africa. This data was tabulated and visualized on a global map using R studio (https://www.r-project.org). A bar representing African YF cases and deaths according to year of occurrence and Country was also generated in R studio (https://www.r-project.org). The methods employed in this report has been published as a preprint in MedRxiv (https://www.medrxiv.org/node/690912.full).

### Phylogenetic analysis

Partial genome sequences downloaded from GenBank were aligned using MAFFTv7.222 (FF- NS-2 algorithm) following default settings [18]. The dataset consisted genome sequences from Africa available in GenBank as at 1^st^ February 2023, along with representative sequences from the Southern America (n =220). Maximum likelihood phylogenetic analysis was performed using the general time reversible nucleotide substitution model after automatic model testing with gamma distributed rate variation GTR-Γ[19] with 1000 bootstrap replicates using IQ-TREE software [20]. The pairwise amino acid distances of the African YF sequences were analyzed in reference to the prototype vaccine Asibi strain (GenBank no: AY640589.1) using MEGA 7 (21).

### Phylogeography and Evolutionary analysis

Evolutionary and additional phylogenetic analysis was carried out on African YF strains, using a Bayesian evolutionary approach using Markov Chain Monte Carlo (MCMC) implemented in BEAST version 2.5 [22]. The strength of the temporal clock signal of the dataset was carried out by conducting a regression of root to tip genetic distances against year of sampling analysis using TempEst v 1.5 [23]. For the MCMC run, a total of 110 YF virus partial E gene sequences of 280 nucleotides in length were aligned, consisting 91 African sequences and 19 Chinese sequences from the international outbreak in 2016. A list of the sequences used for the analysis is contained in supplementary table 2. Further analysis was then done using the strict clock with coalescent Bayesian Skyline prior. The MCMC run was set at 100,000,000 states with a 10% burn in. Results were visualized in Tracer version 1.8. (http://tree.bio.ed.ac.uk/software/tracer/).

Bayesian skyride analysis was carried out to visualize the epidemic evolutionary history using Tracer v 1.8. (http://tree.bio.ed.ac.uk/software/tracer/).

Geographical coordinates of the locations of the African YF sequences were retrieved from the web with the help of online servers. A phylogeographic tree with discrete traits was constructed using the African YF sequences and their geographic coordinates in latitude and longitude using a Bayesian stochastic search variable selection (BSSVS) model implemented in BEAST 2.5 [22]. The resulting tree was annotated in TreeAnnotator after discarding a 10% burn-in. In addition, a continuous phylogeographic tree was also constructed for major African YF genotypes, according to their regions of circulation, WA-I/II, and EA-1/EA-II-Angolan genotypes, using a similar approach for the discrete tree except that a continuous tree trait was selected in creating the BEAST XML file in Beauti. The resulting MCC trees were visualized in ggtree [24].

### Geographic dispersal

The transmission histories and geographic dispersal was projected using the Phylogeographic trees (Continuous and Discrete) were loaded unto an online software sever where information such as internal node ages, user determined geographic coordinates e.t.c. and maps these to a world geographic map. rendered into SPREAD4 [25].

## RESULTS

### Yellow fever outbreak Demography

African Yellow fever demography was re-evaluated with updated data from recent literature [13, 26]. There were also silent outbreaks identified from molecular detection from patients with pyrexia of unknown origin in Nigeria [16]. From our analysis, Nigeria remains the country with highest number of cases, with Ethiopia having the least number of cases (Figure 2 top). The case fatality distribution shows that high mortality rates were recorded between 1984 and 1987 Figure 2 bottom, with majority of cases arising from Nigeria, Angola and Ghana Supplementary table 1.

**Figure 2.**
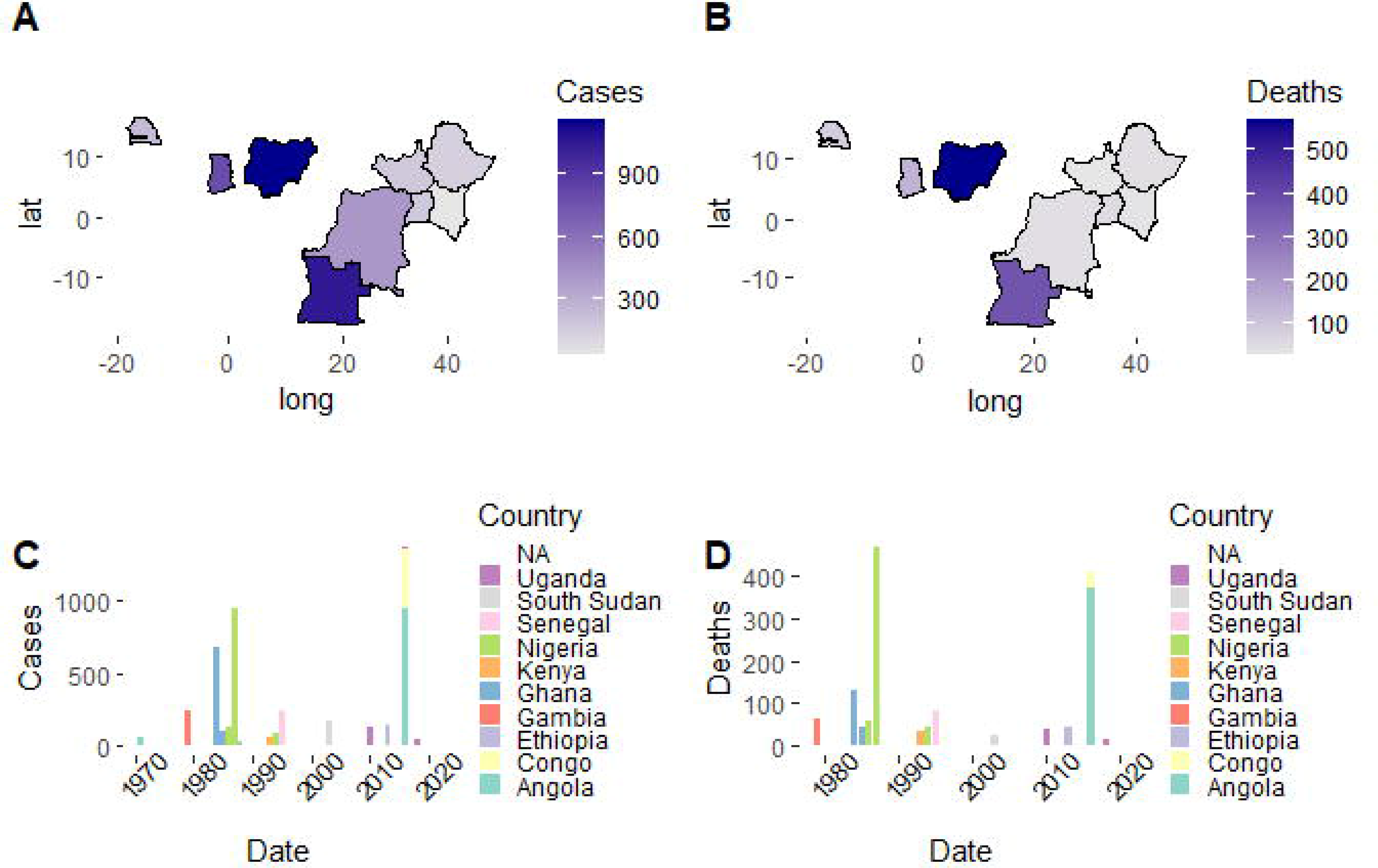
A Geographic distribution of YF virus cases across Africa, color intensity is representative of the number of confirmed YF cases in each country and is displayed as a color gradient by the side of the map. Figure 2B Geographic distribution of YF virus deaths in Africa, color intensity is representative of the number of confirmed YF deaths in each country and is displayed as a color gradient. Figure 2C Bar charts showing the number of confirmed Yellow Fever cases from 1970 to 2021 according to Country. The legend is color coded to represent the various countries where outbreaks occurred. **2D** Bar charts showing the number of confirmed Yellow Fever deaths from 1970 to 2021 according to Country. The legend is color coded to represent the various countries where outbreaks occurred.

### Phylogeography and Evolution

Global phylogeny of YF virus revealed that the African viruses clustered within the four major genotypes West African genotype I (WA-I), West African genotype II (WA-II), East African genotype (EA), Angolan genotype or East African genotype II (EA-II). Majority of the African strains fell under the WA-II, while WA-I had the lowest number of viral sequences as shown in Figure 3. There was strict geographic defined phylogenetic diversification as shown by continental clustering of the viruses analyzed in our study with the South American strains clustering within the South American genotype and African strains within the African genotypes. The amino acid divergence of the African YF partial E gene sequences ranged between 0% to 5.9%, with the recently isolated West African genotypes displaying very low diversity, while the older East African genotypes having between 1.2% to 5.9% (Supplementary table 3).

**Figure 3.**
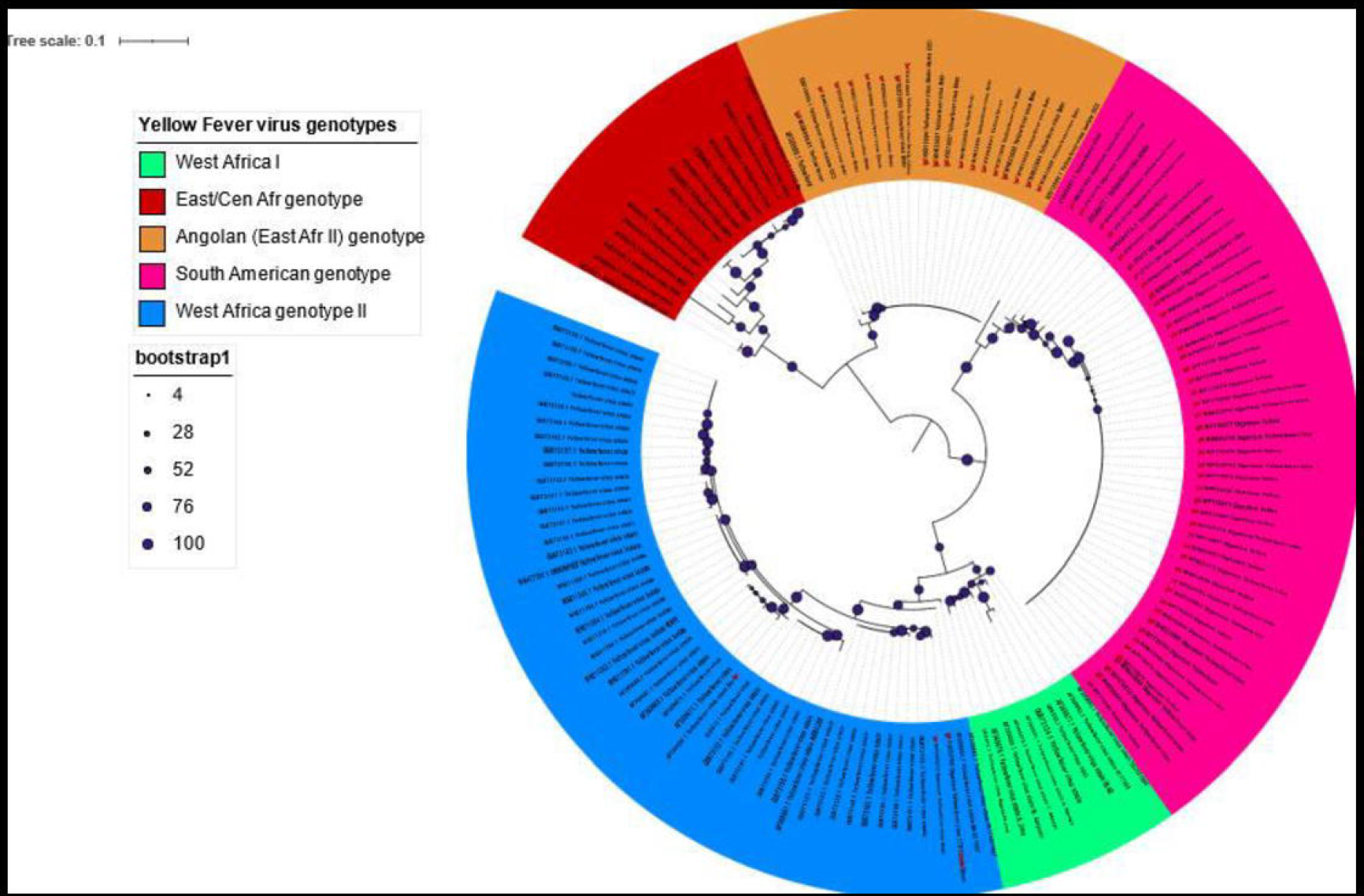
Maximum Likelihood tree of African Yellow Fever isolates along with some global reference sequences. The clades representing the various genotypes are shown as colored rings, the bootstrap valuses are shown as circles around the ancestral nodes with the sizes if the circles representing the percentage bootstap values. The root of the tree is at the mid-point. The legend represents the various global YF genotypes.

The root to tip divergence for the African YF sequences showed a positive signal, with *R^2^*= 0.11 (Figure 4A). The MCC tree of the African YF isolates showed a similar diversification with the global tree with Senegalese isolates being the most abundant and diversly distributed within both WA-I and WA-II genotype sub-clades (Figure 4 bottom). The evolutionary rate of the African YF viruses was 2.08 × 10^-4^, 98% HPD (8.48 × 10^-5^ - 3.41 × 10^-3^) substitutions/ site/ year. The time calibrated MCC tree also showed ancestral divergence times for each ancestral node with the earliest ancestor dating back as the year 1140. The WA-I genotype had a time to most recent ancestor TMRCA of 1845, 95% HPD (1795 – 1890), while WA-II had a TMRCA of 1852, 95% HPD (1790 – 1910). The EA genotype on the other hand had a TMRCA of 1760, 95% HPD (1655 – 1850), and EA-II 1940, 95% HPD (1900 – 1970). Other ancestral nodes for the genotypes, particularly WA genotypes displayed high posterior support. The viral population of the African sequences had no population growth between 1930 to 1970 but exhibited a state of equilibrium, a sharp decline in effective viral population was however observed between 1980 to the most recent sampling date (Supplementary Figure 1).

**Figure 4.**
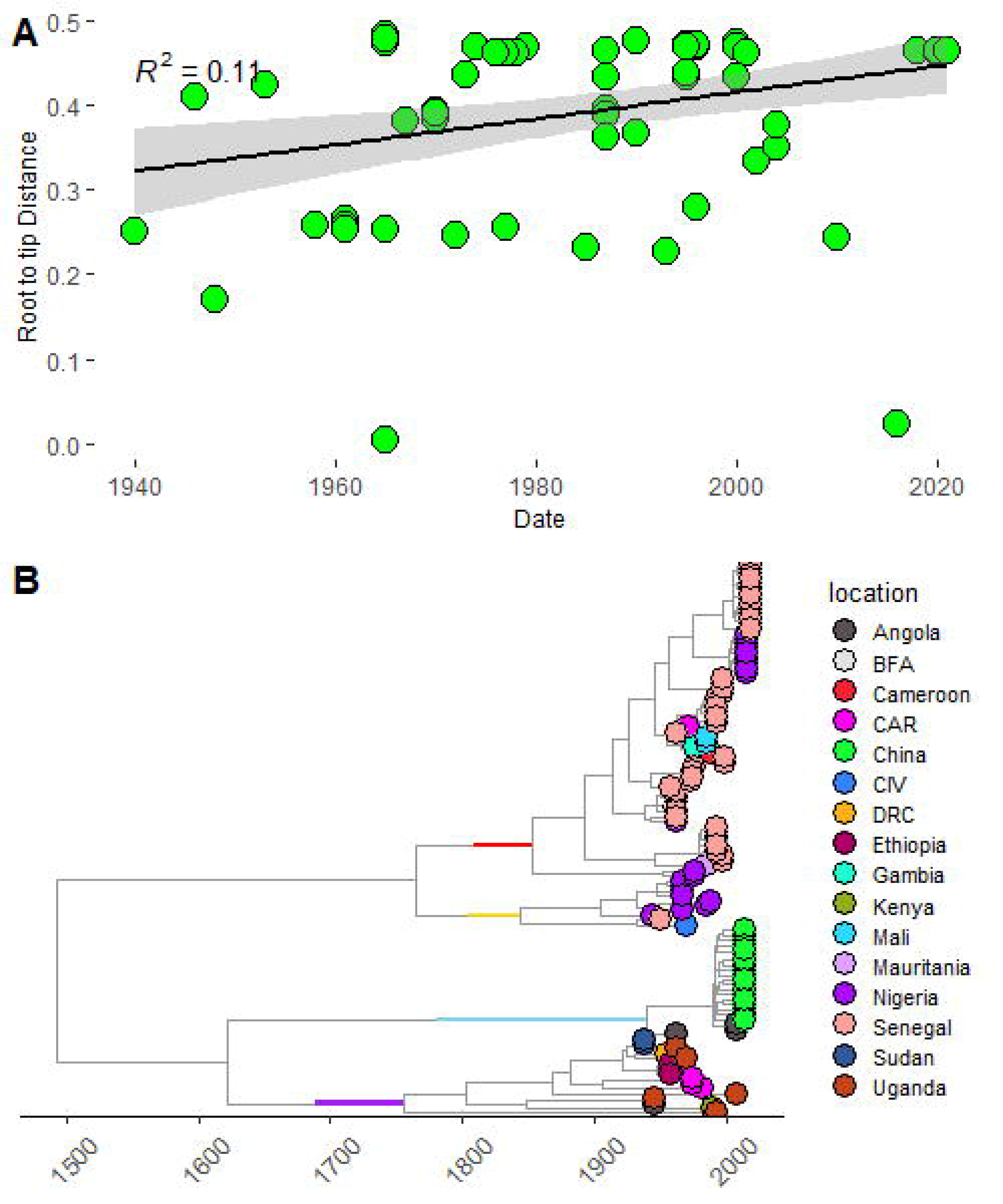
A. Root to tip regression analysis of African Yellow Fever sequences, represented by a scatterplot with a regression line of best fit, shaded portion of the line is the 95% posterior probability. **4B** Time resolved phylogeographic tree of African Yellow virus E gene sequences, showing the various Countries of origin as tips. The genotypes are represented by colors bars at the ancestral nodes of each genotype in the tree. The red bar represents West Africa genotype II (WAII), the yellow bar represents WA, the sky-blue bar represents the Angolan genotype, while the purple bar represents the East African genotype (EA).

### Yellow fever virus geographic dispersal

We explored the pattern of geographic dispersal and viral circulation of YF virus within Africa using both discrete and continuous phylogeographic reconstructions. The discrete model clearly showed major transmission routes between East and West Africa. West African countries of Senegal, Burkina Fasso, and Mauritania all seemed to have outbound transmission linkages to other countries, majorly in East and Central Africa. The strongest transmission links visualized as the width of the connecting cords were seen between, Senegal and DRC, Cote d’Ivoire and Angola, and Mauritania and Angola (Figure 5). The phylogeographic movement seemed to be predominantly from West to East Africa, with only few transition events moving out of East Africa to West African countries. For our continuous phylogeography we ran independent analysis on each the two prevailing genotypes of West Africa and East Africa, giving us two independent results. Our analysis was designed to identify specific viral introductions and spread events from historically endemic areas through time (Figure 6A and 6B). In Figure 6A the continuous spatial dispersion of YF West African genotypes (WAI and WAII), shows that between 1960 and 1980, YF fever spread from Senegal and Mali eastwards into countries such as Nigeria, Ghana, and as far as DRC (Figure 6A). There was also transmission from Nigeria eastwards into Cote d’Ivoire showing some eastward movement as well. The most recently observed transmission route was however, from Ghana into Cameroon and from Ghana back into Nigeria occurring between 1980 to 2021. For the East African genotypes shown in Figure 6B, the epicenter of the EA YF spread was around the DRC and between 1750 and 1910, the genotype was spatially dispersed into majorly East and Central African countries such as Angola, Uganda and Kenya. The most recent special movement of the EA genotype were upward from the DRC into Ethiopia and Sudan, between 1910 and 2010.

**Figure 5.**
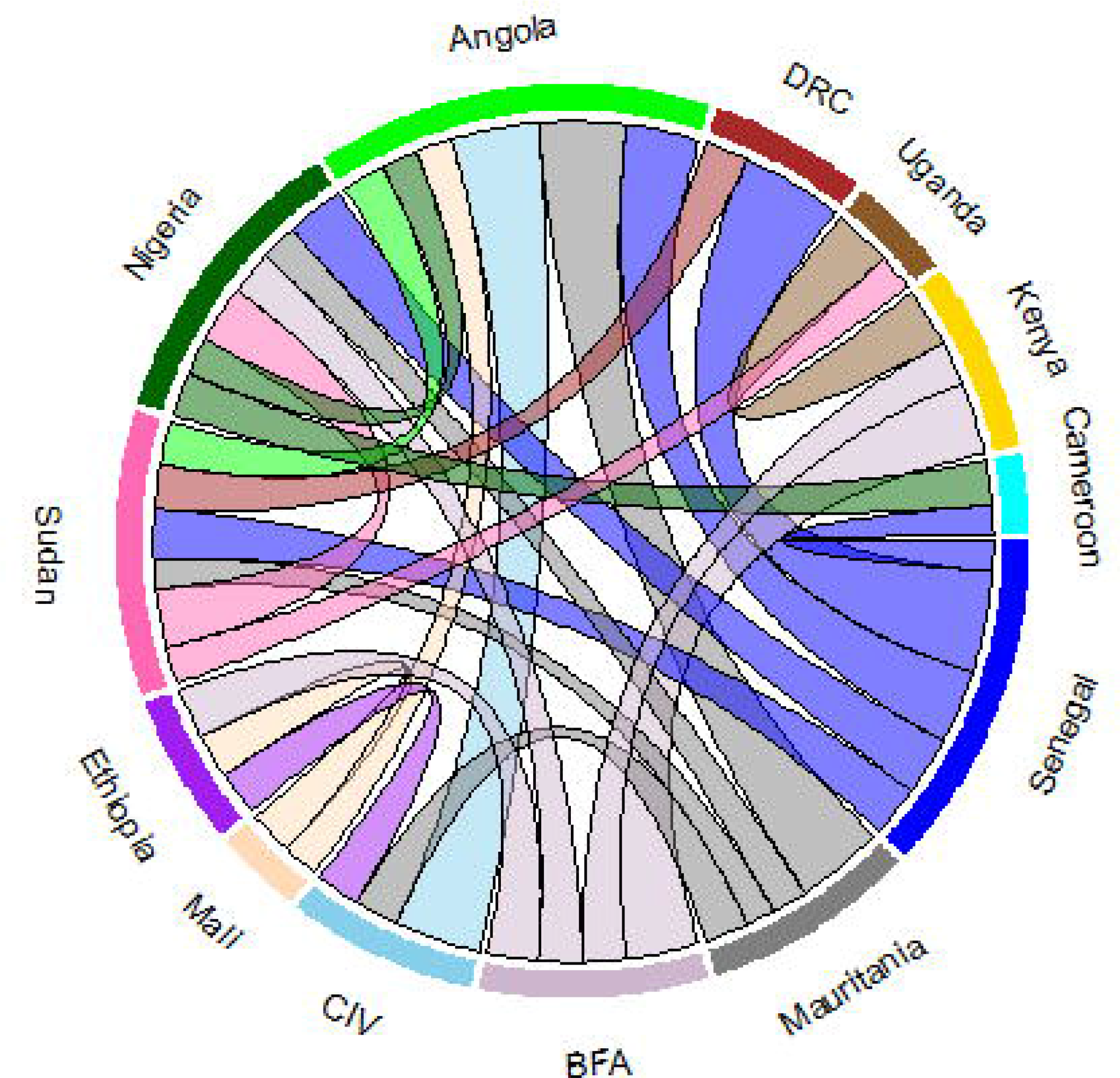
Spatiotemporal diffusion with discrete traits of Yellow Fever virus in Africa after phylogeographic analysis, visualized as a chord plot with the thickness of the connecting chords representing the strength of the geographic linkages between the countries in Bayes Factor (BF).

**Figure 6.**
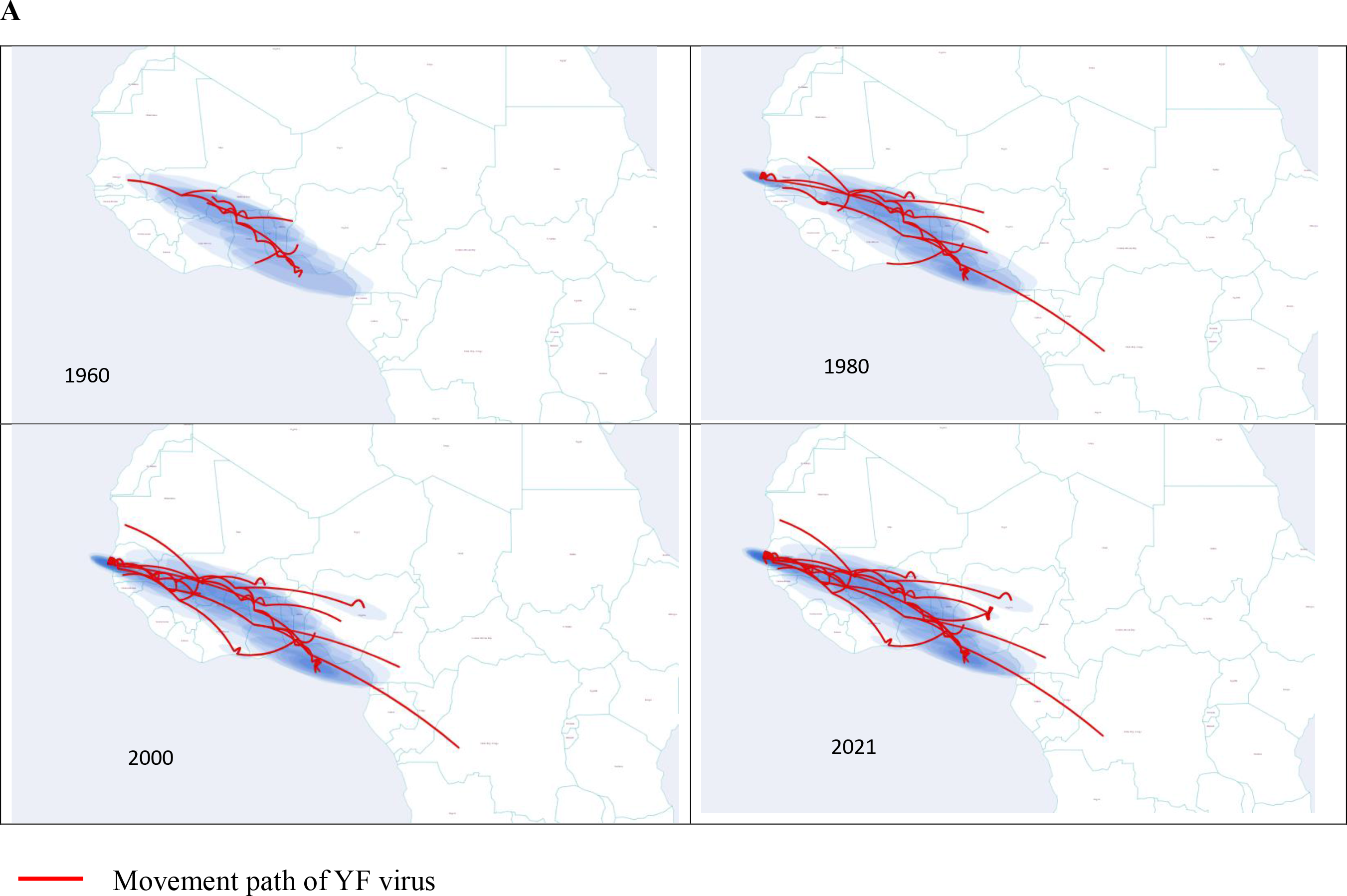
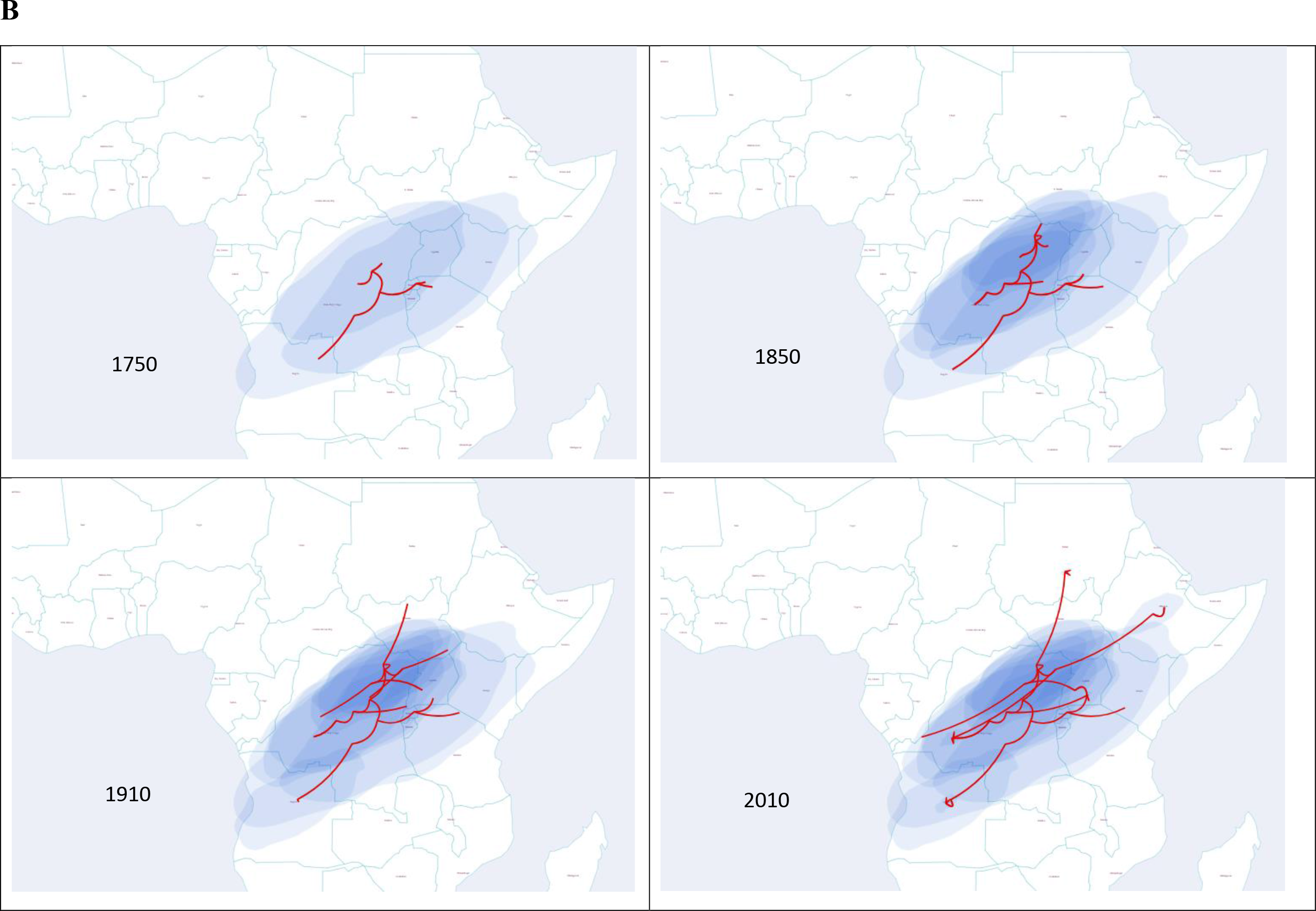
A. Continuous phylogeography of West African Yellow Fever genotypes, shaded blue circles represent 80% highest posterior density interval of the respective nodes of the MCC tree. Directionality of the movements is downward curve left to rightward movement; upward curve is right to leftward movement. **6B**. Continuous phylogeography of East African/Angolan Yellow Fever genotypes, shaded blue circles represent 80% highest posterior density interval of the respective nodes of the MCC tree.

## DISSCUSION

### General and Molecular Epidemiology

Yellow fever virus was first reported in America around the 15^th^ century by Spanish conquerors, but historical and molecular evidence indicates that the virus first originated in Africa where it was imported into the Americas through the trans-Atlantic slave trade [10, 27], ever since then there has been several large outbreaks in mostly West Africa and parts of Central Africa [28].

Yellow fever virus was first isolated from the blood of a Ghanian man called Asibi in 1927 [29]. Epidemiological data from previous outbreaks, between 1960 and 2022 reviewed in our study indicates a fluctuating pattern of intermittent outbreaks with its peak at between 1986 and 1988 (Figure 1b). There was also an upsurge in cases resulting from a large outbreak in Angola in 2015, which spilled over into the Democratic Republic of Congo, followed by Uganda 2016. It is presumed that certain social drivers were largely responsible for this re-emergence of large epidemics in East Africa, these include long term conflict among waring rebels and government forces, human population displacement resulting from conflicts, and improved surveillance activity.

Molecular epidemiology of the YF virus was consistent with previous reports [6, 30, 31]. With the African sequences maintaining their geographically defined genotypes (Figure 3). There was one West African genotype II reported from the Central African Republic GenBank accession number GU073130. The virus was isolated from a mosquito in 1974 in Central African Republic, an indication that the WA-II genotype might have been circulating and maintained in the forests of CAR. The EA genotypes I and II were strictly maintained in their geographical region. The majority of the recent sequences were of WA-II, with the genotype dominating the Entire West Africa and parts of Central Africa. The evolutionary rate of 2.08 X 10^-4^ is lower that of a previous report in Africa of 2.8 × 10-4 [11]. Our result is also lower than that of a study of Brazilian YF isolates of 3.1 × 10-4 [32], but is consistent with that of Sall et al [30] which gave 2.10 × 10-4. This shows that the possible mutation rate of YF virus has remained fairly stable through the years in Africa, and is probably slower than isolates causing outbreaks in South Africa.

### Phylogeography and transmission dynamics

There is a paucity of data regarding special epidemiology of Infectious agents in Africa, recent reports from West Africa have shown the significance of molecular biogeography and special epidemiology in tracking rapidly expanding infectious agents within West Africa [33–35].

Yellow fever phylogeography have consistently showed a distinct geographically bound diversification [6, 30, 36]. Our present study maintains the same regional clustering of the West African genotypes and the East/Central African genotype as well as the Angolan genotype. The more recent sequences isolated from Nigeria in 2018, clustered within WA-II, having a common ancestral history as other closely related isolates, sharing the same sub-clade with isolates from Senegal, Cameroon, and Burkina Fasso. This observation shows strict geographical conservation probably due to the fact that same genotype has established itself within the Forests and grasslands of Nigeria all the way to Senegal. Previous studies have hypothesized that sustained epizootic transmission cycles within forested regions of Africa have led to YF virus adaptation to its intermediate host NHP and its competent vector *Heamagogous spp* [6]. Also, the fact that humans act as a dead-end host leaves little opportunity for intra-host evolution and diversity among human population. Previous reports have identified key mutational features among African YF isolates that confer a positive selection pressure and may play a role in genotype emergence [10, 36]. The EA-I and the EA-II/Angolan genotypes were in conformity with previously reported geographically defined diversity [6, 30]. The most recently identified strains from outbreak in Angola and Uganda 2015/2016 clustered within separate genotypes.

Discrete phylogeography showed active transmission routes within both West Africa and East Africa and between both regions. This is an indication of sustained intra-regional transmission as well as trans regional transmission resulting from constant human movement between West and Central Africa. This observation serves as evidence that YF transmission dynamics is not just influenced by ecological drivers and climatic events that may affect the Forest /Savanah ecosystem as previously reported [37–39], but also by human activity such as international trade and human migration. Continuous phylogeography revealed a gradual temporal spread among the genotypes within their regions of primary emergence. The WA genotypes seemed to have stronger migration events compared to the EA genotypes. The WA genotypes displayed a eastward movement cutting across more than 4 countries within the space of just 40 years from Senegal, Mali axis all the way to Camerron as far east as Angola. This very fast spread of the WA genotype may not be unconnected with the rich vegetation across the region and the successful adaptation of the principal insect vector *aedes egypti* to the urban environment of the cities of West Africa. The EA genotype however displayed a much slower spread rate, with its spread event mainly localized within the Central/East African regions. Although there were very recent introductions northward to Sudan and Ethiopia, this seems to have occurred as isolated incidents. Our results in this study affirms that vegetation plays a vital role in the geographic spread of YF across Africa as there was limited Northward geographic movement of the WA genotypes (Figure 6a). The EA/Angolan genotypes seemed to be restricted in their dispersal activity within the East/Central African region, although some northward movement was observed with spread events into Sudan and Ethiopia, these were just probably isolated occurrences arising from local outbreaks from Sylvatic/Sahvana YF cycle spillovers into human habitation areas.

## CONCLUSION

This study reports the most current data on YF virus phylogeography and transmission dynamics. We have shown that YF virus maintain a strict geographically bound lineage dispersal pattern, with major genotypes circulating within strict regional confines. We also report a major surge in reported laboratory confirmed cases majorly in East Africa, after the year 2015. There was also rapid expansion of the WA-II genotype in the last 4 years. Study limitations include poor genomic surveillance system across Africa, resulting in limited amount of YF sequence data available in open-source data banks. We therefore recommend expanded human and vectoral surveillance of YF outbreaks as well as other Arboviruses of public health importance. We also advocate for upscaling Molecular biology capabilities of existing public health labs both Nationally and regionally to enable rapid genomic sequencing and analysis of samples from Arboviral outbreaks such as YF to help in prevention of future epidemics and protect Africa from existing biological threats.

## DATA AVAILABILITY STATEMENT

The data used in this study is freely available on GenBank and their information is also available as a supplementary table.

## REFERENCES

1. Monath TP, Vasconcelos PFC. Yellow fever. Journal of Clinical Virology. 2015;64: 160–173. 10.1016/j.jcv.2014.08.030.

2. Beasley D.W., McAuley A.J., Bente D.A. Yellow fever virus: Genetic and phenotypic diversity and implications for detection, prevention and therapy. Antivir. Res. 2015;115:48–70. doi: 10.1016/j.antiviral.2014.12.010.

3. Gould EA, de Lamballerie X, Zanotto PM, Holmes EC. 2003. Origins, evolution, and vector/host coadaptations within the genus Flavivirus. Adv. Virus Res. 59:277–314.

4. Vasconcelos PFC, Sperb AF, Monteiro HAO, Torres MAN, Sousa MRS, Vasconcelos HB, Mardini LB, Rodriguez SG. Isolations of yellow fever virus from Haemagogus leucocelaenus in Rio Grande do Sul State, Brazil. Transactions of the Royal Society of Tropical Medicine and Hygiene. 2003; 97: 60–62.

5. Chang, G. J., Cropp, B. C., Kinney, R. M., Trent, D. W., & Gubler, D. J. Nucleotide sequence variation of the envelope protein gene identifies two distinct genotypes of yellow fever virus. Journal of virology, 1995.69(9), 5773–5780.

6. Mutebi, J. P., Wang, H., Li, L., Bryant, J. E., & Barrett, A. D. Phylogenetic and evolutionary relationships among yellow fever virus isolates in Africa. Journal of virology, 2001.75(15), 6999–7008. doi:10.1128/JVI.75.15.6999-7008.2001.

7. Siddle, K. J., Eromon, P., Barnes, K. G., Mehta, S., Oguzie, J. U., Odia, I., Schaffner, S. F., Winnicki, S. M., Shah, R. R., Qu, J., Wohl, S., Brehio, P., Iruolagbe, C., Aiyepada, J., Uyigue, E., Akhilomen, P., Okonofua, G., Ye, S., Kayode, T., Ajogbasile, F., … Happi, C. T. Genomic Analysis of Lassa Virus during an Increase in Cases in Nigeria in 2018. The New England journal of medicine, 2018. 379(18), 1745–1753. doi:10.1056/NEJMoa1804498.

8. Carroll, M. W., Matthews, D. A., Hiscox, J. A., Elmore, M. J., Pollakis, G., Rambaut, A., Hewson, R., García-Dorival, I., Bore, J. A., Koundouno, R., Abdellati, S., Afrough, B., Aiyepada, J., Akhilomen, P., Asogun, D., Atkinson, B., Badusche, M., Bah, A., Bate, S., Baumann, J., … Günther, S. Temporal and spatial analysis of the 2014-2015 Ebola virus outbreak in West Africa. Nature,2015. 524(7563), 97–101. doi:10.1038/nature14594.

9. Motayo, B. O., Oluwasemowo, O. O., Olusola, B. A., Opayele, A. V., & Faneye, A. O. Phylogeography and evolutionary analysis of African Rotavirus a genotype G12 reveals district genetic diversification within lineage III. Heliyon, 2019. 5(10), e02680. doi:10.1016/j.heliyon.2019.e02680.

10. Stock NK, Laraway H, Faye O, Diallo M, Niedrig M, Sall AA. 2013. Biological and Phylogenetic Characteristics of Yellow Fever Virus Lineages from West Africa. J Virol 87:10.1128/jvi.01116-12.

11. Beck A, Guzman H, Li L, Ellis B, Tesh RB, Barrett ADT (2013) Phylogeographic Reconstruction of African Yellow Fever Virus Isolates Indicates Recent Simultaneous Dispersal into East and West Africa. PLoS Negl Trop Dis 7(3): e1910.

12. WHO Countries with risk of yellow fever transmission and countries requiring yellow fever vaccination.2020. https://www.who.int/publications/m/item/countries-with-risk-of-yellowfever-transmission-and-countries-requiring-yellow-fever-vaccination-(july-2020).

13. Nomhwange, T., Jean Baptiste, A. E., Ezebilo, O., Oteri, J., Olajide, L., Emelife, K., Hassan, S., Nomhwange, E. R., Adejoh, K., Ireye, F., Nora, E. E., Ningi, A., Bathondeli, B., & Tomori, O. The resurgence of yellow fever outbreaks in Nigeria: a 2-year review 2017-2019. BMC infectious diseases, 2021. 21(1), 1054. 10.1186/s12879-021-06727-y.

14. Nwachukwu WE, Oladejo J, Ofoegbunam CM, Anueyiagu C, Dogunro F, Etiki SO, Dachung BI, Obiekea C. Epidemiological description of and response to a large yellow fever outbreak in Edo state Nigeria, September 2018 - January 2019. BMC Public Health. 2022. 22(1):1644. doi: 10.1186/s12889-022-14043-6.

15. Dieng, I., Diallo, A., Ndiaye, M., Mhamadi, M., Diagne, M. M., Sankhe, S., Ndione, M. H. D., Gaye, A., Sagne, S. N., Heraud, J. M., Sall, A. A., Fall, G., Loucoubar, C., Faye, O., & Faye, O. Full genome analysis of circulating DENV-2 in Senegal reveals a regional diversification into separate clades. Journal of medical virology,2022. 94(11), 5593– 5600.

16. Ajogbasile, F. V., Oguzie, J. U., Oluniyi, P. E., Eromon, P. E., Uwanibe, J. N., Mehta, S. B., Siddle K .J, Odia I, Winnicki S. I, Akpede N, Akpede G, Okogbenin S, Ogbaini-Emovon E, MacInnis B.L, Folarin O.A, Modjarrad K, Schaffner S.F, Tomori O, Ihekweazu C, Sabeti P. C, Happi C. T. Real-time Metagenomic Analysis of Undiagnosed Fever Cases Unveils a Yellow Fever Outbreak in Edo State, Nigeria. Scientific reports, 2020. 10(1), 3180. 10.1038/s41598-020-59880-w.

17. Li C, Li D, Smart S.J, Zhou L, Yang P, Ou J. Evaluating the importation of Yellow Fever cases into China in 2016 and strategies used to prevent and control the spread of the disease. 2020. WPSAR. 11(2): doi: 10.5365/wpsar.2018.9.1.007.

18. Katoh K, Misawa K, Kuma K, Miyata T, MAFFT: a novel method for rapid multiple sequence alignment based on fast Fourier transform, *Nucleic Acids Research*, Volume 30, Issue 14, 15 July 2002, Pages 3059–3066.

19. Yang Z. Estimating the pattern of nucleotide substitution. Journal of molecular evolution, 1994. 39(1), 105–111.

20. Nguyen, L.T.; Schmidt, H.A.; von Haeseler, A.; Minh, B.Q. IQ-TREE: A fast and effective stochastic algorithm for estimating maximum-likelihood phylogenies. Mol. Biol. Evol.2015. 32, 268–274. 10.1093/molbev/msu300

21. Kumar S, Stecher G, and Tamura K. Molecular Biology and Evolution. 2016. 33:1870–1874

22. Bouckaert R., Vaughan T.G., Barido-Sottani J., Duchêne S. BEAST 2.5: An advanced software platform for Bayesian evolutionary analysis. PLoS computational biology, 2019. 15(4), e1006650. doi:10.1371/journal.pcbi.1006650.

23. Rambaut, A. Lam, T.T. Carvalho, L.M. Pybus, O.G. Exploring the temporal structure of heterochronous sequences using TempEst (formerly Path-O-Gen). Virus Evol. 2020. 2, vew007.

24. Yu, G., Smith, D. K., Zhu, H., Guan, Y. & Lam, T.T.-Y. ggtree: An r package for visualization and annotation of phylogenetic trees with their covariates and other associated data. Methods Ecol. Evol. 2017. 8, 28–36.

25. Nahata, K. D., Bielejec, F., Monetta, J., Dellicour, S., Rambaut, A., Suchard, M. A., Baele, G., & Lemey, P. SPREAD 4: online visualisation of pathogen phylogeographic reconstructions. Virus evolution, 2022. 8(2), veac088. 10.1093/ve/veac088.

26. World Health Organization. Yellow fever situation report, 9 June2016. http://www.who.int/emergencies/yellow-fever/situation-reports/9-june-2016/en/

27. White C.R. Yellow fever; history of the disease in the eighteenth and nineteenth century. J. Kans. Med. Soc. 1959. 60:298–302.

28. Chippaux J.P., Chippaux A. Yellow fever in Africa and the Americas: A historical and epidemiological perspective. J. Venom. Anim. Toxins. Incl. Trop. Dis. 2018. 24:20. doi: 10.1186/s40409-018-0162-y.

29. Staples, J. E., & Monath, T. P. (2008). Yellow fever: 100 years of discovery. JAMA, 300(8), 960–962. 10.1001/jama.300.8.960.

30. Sall AA, Faye O, Diallo M, Firth C, Kitchen A, Holmes EC. Yellow fever virus exhibits slower evolutionary dynamics than dengue virus. J Virol. 2010. 84(2):765–772.

31. Grobbelaar, A. A., Weyer, J., Moolla, N., Jansen van Vuren, P., Moises, F., & Paweska, J. T. Resurgence of Yellow Fever in Angola, 2015-2016. Emerging infectious diseases, 2016. *22*(10), 1854–1855.

32. Nunes MRT, Palacios GCardoso JF, Martins LC, Sousa EC, de Lima CPS, Medeiros DBA, Savji NDesai A, Rodrigues SG, Carvalho VL, Lipkin WI, Vasconcelos PFC. 2012. Genomic and Phylogenetic Characterization of Brazilian Yellow Fever Virus Strains. J Virol 86:.10.1128/jvi.00565-12.

33. Motayo B.O, Oluwasemowo O.O, Akinduti P.A. Evolutionary dynamics and geographic dispersal of beta coronaviruses in African bats. PeerJ. 2020. 8:e10434. 26. doi:10.7717/peerj.10434.

34. Akinduti P.A, Ayodele O, Motayo BO, Obafemi YD, Isibor PO, Aboderin OW. Cluster analysis and geospatial mapping of antibiotic resistant *Escherichia coli* O157 in southwest Nigerian communities. One Health. 2022;15:100447. doi:10.1016/j.onehlt.2022.100447.

35. Akinduti P.A, Osiyemi JA, Banjo TT. Clonal diversity and spatial dissemination of multi- antibiotics resistant Staphylococcus aureus pathotypes in Southwest Nigeria. PLoS One. 2021;16(2):e0247013. doi:10.1371/journal.pone.0247013

36. Li Y. Molecular epidemiology of yellow fever virus in Africa: A perspective of the phylogeographic split between East/Central African and West African lineages. Acta tropica, 2022. 225, 106199. 10.1016/j.actatropica.2021.106199.

37. Vasconcelos PF, Bryant JE, da Rosa TP, Tesh RB, Rodrigues SG, Barrett ADT. Genetic divergence and dispersal of yellow fever virus, Brazil. Emerg Infect Dis. 2004. 10: 1578– 1.

38. Moreira-Soto A, Torres MC, Mendonc _a MCL de, Mares-Guia MA, Rodrigues CD dos S, Fabri AA, DosSantos CC, Fisher C, Ribeirio Nogueira RM, Drosten C, Drexler JF, Bispo de Filipos AM. Evidence for multiple sylvatic transmission cycles during the 2016–2017 yellow fever virus outbreak, Brazil. Clinical Microbiology and Infection. 2018; 24: 1019.e1–1019.e4.

39. Hill, SC., de Souza, R., Thézé, J., Claro, I., Aguiar, RS., Abade, L., Santos, FC. FlaviaC,Salles FCS, Rocco IM, Maeda AY, Vasami FG, Plessis L, Silveira PP, Quick J, Guerra JM, Réssio RA, Giovanetti M, Alcantara LCJ, Cirqueira CS, Díaz- Delgado J, Macedo FLL, Timenetsky MST, Mucci LF, Tubaki RM, de Menezes R, Ramos P, Cruz LN, Loman N, Dellicour S, Pybus OG, Ester Sabino EC, Faria NR. Genomic Surveillance of Yellow Fever Virus Epizootic in São Paulo, Brazil, 2016 - 2018. *PLoS pathogens*, 2020. *16*(8), e1008699.10.1371/journal.ppat.1008699

